# COVID-19 mortality in six Caribbean countries during Four Waves of the COVID-19 Pandemic

**DOI:** 10.64898/2026.10.22.26366162

**Authors:** Jacqueline P Duncan, Affette M McCaw-Binns, J Peter Figueroa

## Abstract

**Background:** Mortality associated with COVID-19 in the Caribbean during specific waves and exposure to different variants has not well described, especially for the six most populous English-speaking countries (Bahamas, Barbados, Guyana, Jamaica, Trinidad and Tobago, and St. Lucia), compared to the United Kingdom (UK).

**Methods:** The first four waves of the pandemic, cases and deaths were estimated from epidemiologic curves on Johns Hopkins University Coronavirus Resource Center for the countries of interest from 10-Mar-20 to 12-Aug-22. Crude mortality rates (CMR), case fatality ratios (CFR) and standardized mortality ratios (SMR) were calculated using England as the standard population.

**Findings:** The six countries reported 550,657 cases and 10,259 COVID-19 deaths. First wave Caribbean CFRs (1-3%) were significantly lower than the UK (20%). Thereafter (1-Apr-21 to 31-May-22) Caribbean SMRs exceeded 1 (range: 1.37-6.35) while UK CFRs declined to 2.3% (2^nd^ wave) and 0.2% (3^rd^ wave). Caribbean CFRs were highest with circulation of the Delta variant in the 2^nd^ and 3^rd^ waves. Barbados had the lowest SMR (0.65). COVID-19 vaccine roll-out (since Feb-Mar 2021) first targeted high-risk groups (elderly, health workers, co-morbidities). No Caribbean country achieved the global full vaccination target (70%) by the 4^th^ wave, range: 21% (Jamaica) to 54% (Barbados), compared to 86% for the UK.

**Interpretation:** Early in the COVID-19 pandemic, Caribbean CFRs were lower than the UK, probably due to more intensive non-pharmaceutical interventions. Limited vaccine coverage negatively impacted mortality in later waves, which was highest when the Delta variant was prevalent, declining with the later Omicron variant.

## Introduction

Coronavirus disease 2019 (COVID-19) has had an unprecedented impact on health care systems, economies and populations worldwide.^1,2^ In countries with high development index (HDI) such as United Kingdom and Italy, high case fatality rates (CFR) and crude mortality rates (CMR) were documented early in the pandemic including CFR>50% for patients requiring invasive mechanical ventilation.^3,4^ Studies confirmed that severe disease and death are associated with older age, male sex, obesity, and underlying conditions such as diabetes mellitus, hypertension and heart failure.^3,5^ Successive waves in HDI countries following Alpha and Delta dominant strains showed declines in CFRs due to vaccination roll out, treatment availability, increased testing, less virulent circulating variants of concern (VOC), natural immunity as well as improved management based on better understanding of the disease process.^6^

Early in the pandemic, many low and middle income countries (LMIC) including African and Caribbean countries were thought to be less affected by COVID-19 due to CFRs of 1 to 3% compared to >10% in high income countries (HIC).^7^ This spurned speculations about protective factors such as BCG vaccine and sunlight.^8,9^ However, studies failed to show any significant association between these factors and severe COVID-19. Analyses of COVID-19 cases and deaths in African countries in subsequent waves showed higher CFR than the global level (CFR >2.2%).^10^

As of 12-Sep-2022, 4,216,157 COVID-19 cases and 35,445 COVID-19 deaths were reported across 35 countries in the Caribbean.^11^

The Caribbean Public Health Agency (CARPHA) reports a CFR of <1.0% for the Caribbean as of 12-Sep-2022.^11^ However, these data are from settings with limited testing and varying methods of case ascertainment. Case fatality rates (CFR) of 0 to 9.1% have been reported among children 0 to 19 years in Latin America and the Caribbean (LAC).^12^ Among children 0 to 9 years old, CFRs ranging from 0.0% in Jamaica to 2.8% in Haiti were recorded in 2021.^13^ Differences in CMRs and CFRs between countries during separate waves of COVID-19 are important to better understand the epidemiology of COVID-19. However the pattern of COVID-19 mortality across the Caribbean, including mortality during waves with different variants of concern (VOC), is not adequately described.. These rates can be influenced by many factors including ascertainment of cases and deaths due to COVID-19, population age structure, population density, levels of testing, compliance with non-pharmaceutical interventions, vaccination coverage, treatment availability and virulence of the circulating VOC.

Standardization is one method to control for variations in age distribution when comparing mortality rates between countries. The aim of our study is to describe the pattern of COVID-19 mortality in the six most populous English-speaking Caribbean countries. We estimated the CMR and CFR during the first, second, third and fourth waves as well as the standardized mortality ratio (SMR) over 1-year periods using a high-income country, England, as the standard population.

## Materials and Methods

The number of COVID-19 cases and deaths up to 12-Aug-2022 were determined for the United Kingdom (UK) and the six most populous English-speaking Caribbean countries (by population): Jamaica, Trinidad and Tobago (T&T), Guyana, Bahamas, Barbados and St. Lucia. The epidemiologic curves for daily reported COVID-19 cases and deaths for these Caribbean countries and the UK were reviewed on the Johns Hopkins University Coronavirus Resource Center.^14^

### COVID-19 waves and case ascertainment

The plots of COVID-19 cases and deaths by date (month, day, year) were used to estimate the start (rapid increase in cases) and end dates (return to lowest 7-day average between waves) for the first (1^st^), second (2^nd^), third (3^rd^) and fourth (4^th^) waves of COVID-19 for each country (Table 1). We confirmed the approximate start and end times for each wave by reviewing CARPHA and Ministry of Health announcements as well as reports from local news media.

**Table 1:** Estimated start and end dates of first, second, third and fourth waves of COVID-19 in six Caribbean countries and UK.

| **Country** | **1st wave** | **2nd wave** | **3rd wave** | **4th wave** |
| --- | --- | --- | --- | --- |
| **Bahamas** | 15-Mar-20 to 4-Dec-20 | 9-Mar-21 to 7-Jun-21 | 8-Jun-21 to 12-Nov-21 | 6-Dec-21 to 8-Mar-22 |
| **Barbados** | 17-Mar-20 to 10-May-21 | 19-Aug-21 to 17-Dec-21 | 18-Dec-21 to 24-Mar-22 | 23-Mar-22 to 17-Jun-22 |
| **Guyana** | 11-Mar-20 to 28-Dec-20 | 28-Dec-20 to 30-Jul-21 | 31-July-21 to 25-Dec-21 | 26-Dec-21 to 17-Mar-22 |
| **Jamaica** | 9-Mar-20 to 30-Dec-20 | 1-Jan-21 to 1-Jul-21 | 11-Jul-21 to 11-Dec-21 | 17-Dec-21 to 7-Mar-22 |
| **St. Lucia** | 13-Mar-20 to 11-Apr-21 | 21-Jul-21 to 5-Dec-21 | 19-Dec-21 to 29-Mar-22 | 8-Apr-22 to 20-Jun-22 |
| **Trinidad & Tobago** | 12-Mar-20 to 29-Dec-20 | 13-Mar-21 to 3-Aug-21 | 4-Aug-21 to 3-Apr-22 | 4-Apr-22 to 5-Jul-22 |
| **United Kingdom** | 31-Jan-20 to 9-Jun-20 | 10-Jun-20 to 12-Apr-21 | 13-Apr-21 to 24-Feb-22 | 25-Feb-22 to 1-Jun-22 |

The number of COVID-19 cases, and deaths for each wave was determined by taking from country plots, the difference between the cumulative number of COVID-19 cases (or deaths) at the end date of each country’s specific wave from the cumulative number of COVID-19 cases (or deaths) at the start date of their wave. Population size for each country (including age disaggregated data) was determined from The World Bank and *PopulationPyramid.net* databases.^15,16^ Vaccination data were obtained from the *Our World in Data* database and the UK Health Security Agency website.^17,18^ The average of the vaccination coverage at the start and end of each wave was recorded.

### Crude mortality rates (CMR) and case fatality ratios (CFR)

Crude mortality rates were calculated from the total number of COVID-19 deaths in each country for the specified time period divided by the mean total population of the given country, multiplied by 100,000. Case fatality ratios were calculated for each wave by dividing the number of COVID-19 deaths in the specified wave by the number of COVID-19 cases in that wave, multiplied by 100.

### Standardized mortality ratios (SMR)

Indirect standardization using age-specific mortality rates from England was used to determine SMR since the number of deaths by age group was not available for most countries, including Scotland, Wales and Northern Ireland. The number of COVID-19 deaths in England by age group were obtained from reports of the UK Health Security Agency for three time periods: 1) 29-Jun-20 to 31-May-21 and 2) 1-Apr-21 to 31-Mar-22 and 3) 1-Mar-20 and 27-Mar-22. ^19^ These time periods coincide with the availability of final data on annualized mortality rates for England. Age-specific mortality rates were calculated by dividing the number of deaths in laboratory confirmed COVID-19 cases in each age group by the population in that age group.

We applied the age-specific mortality rates for England to the age disaggregated populations (<5, 5 to 9 years, 10 to 19 years, 20 to 29 years etc.) in six Caribbean countries (Bahamas, Barbados, Guyana, Jamaica, St Lucia and Trinidad and Tobago) to estimate expected deaths in each country for the same time periods, had they experienced the same mortality rates as persons of the same age in England. Total expected deaths are the sum of expected deaths for each age group.

Observed deaths for the Caribbean countries for the periods that British data were available were determined from the Johns Hopkins Resource Center graph of cumulative number of deaths against date for each country.^14^ SMRs were calculated by dividing observed number of deaths by expected number of deaths for each country.

### Ethical considerations

We used secondary data on COVID-19 cases and deaths by country for this analysis, so informed consent and ethical approval were not required. The data are de-identified, aggregated and publicly available on a web-based portal.

## Results

Between 1-Mar-2020 and 12-Aug-2022 a total of 550,657 COVID-19 cases and 10,259 COVID-19 deaths were reported among the six most populous English-speaking countries in the Caribbean. The reported cumulative crude mortality rates (CMR) ranged from a low of 108 (Jamaica) to a high of 290 (Trinidad &Tobago) deaths per 100,000 population (Table 2).

**Table 2:** COVID-19 cases and deaths, six Caribbean countries and the UK, 12-Aug-2022.

| **Country** | **Estimated population (2021)** | **Total reported cases** | **Deaths** | **Cumulative cases per 100,000 population** | **Cumulative deaths per 100,000 population** | **Population density per square kilometre** |
| --- | --- | --- | --- | --- | --- | --- |
| Bahamas | 396 914 | 36 850 | 822 | 9 284 | 207 | 38.53 |
| Barbados | 287 708 | 94 963 | 501 | 33 007 | 174 | 666.10 |
| Guyana | 790 329 | 70 159 | 1 272 | 8 877 | 161 | 3.90 |
| Jamaica | 2 973 462 | 147 316 | 3 207 | 4 954 | 108 | 270.99 |
| St. Lucia | 184 401 | 28 161 | 387 | 15 272 | 210 | 298.10 |
| T&T | 1 403 374 | 173 208 | 4 070 | 12 342 | 290 | 270.93 |
| UK | 67 326 569 | 23 634 561 | 205 720 | 35 104 | 306 | 274.70 |

T&T = Trinidad and Tobago
Sources: Our World in Data; Johns Hopkins University Coronavirus Resource Center; Countries By Population Density - WorldAtlas

Using England as the standard population, standardized mortality ratios (SMR) <1 were computed for all countries between 29-Jun-2020 and 31-May-2021 with Barbados having the lowest SMR (0.02) (Table 3). By the second period (1-Apr-2021 to 31-Mar-2021) however, SMRs exceeding one (range: 1.56 to 6.35) were observed for all Caribbean countries, with the highest SMRs noted for T&T (6.35), Bahamas (4.38) and Guyana (3.91). For the reporting period 1-Mar-2020 to 27-Mar-2022 Barbados had the lowest SMR (0.62).

**Table 3:** Standardized mortality ratios (SMR) for six Caribbean countries, 29-Jun-2020 to 31-May-2021 and 1-Apr-2021 to 31-May-2022, using England as the standard population.

|  | **Reporting Period:**<br>**29-Jun-2020 to 31-May-2021** |  |  | **Reporting Period:**<br>**1-Apr-2021 to 31-Mar-2022** |  |  | **Reporting period:**<br>**1-Mar-2020 to 27-Mar-2022** |  |  |
| --- | --- | --- | --- | --- | --- | --- | --- | --- | --- |
|  | **Observed**<br>**Deaths** | **Expected**<br>**Deaths** | **SMR** | **Observed**<br>**Deaths** | **Expected**<br>**Deaths** | **SMR** | **Observed**<br>**Deaths** | **Expected**<br>**Deaths** | **SMR** |
| Bahamas | 219 | 249 | 0.88 | 600 | 137 | 4.38 | 788 | 348 | 2.27 |
| Barbados | 7 | 420 | 0.02 | 332 | 212 | 1.56 | 330 | 536 | 0.62 |
| Guyana | 376 | 469 | 0.80 | 993 | 254 | 3.91 | 1225 | 646 | 1.90 |
| Jamaica | 938 | 2296 | 0.41 | 2289 | 1215 | 1.89 | 2879 | 3042 | 0.95 |
| St. Lucia | 79 | 166 | 0.48 | 310 | 87 | 3.56 | 365 | 218 | 1.68 |
| T&T | 487 | 1050 | 0.46 | 3605 | 567 | 6.35 | 3734 | 1434 | 2.60 |

T&T = Trinidad and Tobago

The first wave of the COVID-19 pandemic in these six Caribbean countries began at similar times (mid-March 2020) and peaked in the second half of 2020 (see Table 1). The UK experienced its first wave between January and June 2020 and had an estimated CFR of 20.2% compared to CFR ranging from 1.1 to 2.6% during the first wave in these Caribbean countries (Table 4).

**Table 4:** Estimated COVID-19 deaths, case fatality rates (CFR), Crude mortality ratios (CMR) and vaccine coverage over four COVID-19 waves, six Caribbean countries and the United Kingdom.

| First Wave |  |  |  |  |  | Second Wave |  |  |  |  | Third Wave |  |  |  |  | Fourth Wave |  |  |  |  |
| --- | --- | --- | --- | --- | --- | --- | --- | --- | --- | --- | --- | --- | --- | --- | --- | --- | --- | --- | --- | --- |
| Country | Deaths | CFR (%) | CMR<sup>A</sup> | Vaccine(%) |  | Deaths | CFR (%) | CMR<sup>A</sup> | Vaccine (%) |  | Deaths | CFR (%) | CMR<sup>A</sup> | Vaccine(%) |  | Deaths | CFR (%) | CMR<sup>A</sup> | Vaccine(%) |  |
|  |  |  |  | Full<sup>B</sup> | ≥1 dose<sup>C</sup> |  |  |  | Full<sup>B</sup> | ≥1 dose<sup>C</sup> |  |  |  | Full<sup>B</sup> | ≥1 dose<sup>C</sup> |  |  |  | Full<sup>B</sup> | ≥1 dose<sup>C</sup> |
| **Bahamas** | 163 | 2.2 | 41 | 0 | 0 | 47 | 1.4 | 12 | 2 | 6 | 433 | 4.1 | 109 | 18 | 23 | 66 | 0.6 | 17 | 37 | 39 |
| **Barbados** | 45 | 1.1 | 16 | 6 | 14 | 208 | 0.9 | 72 | 41 | 46 | 73 | 0.2 | 25 | 52 | 56 | 144 | 0.6 | 50 | 54 | 58 |
| **Guyana** | 164 | 2.6 | 21 | 0 | 0 | 371 | 2.3 | 47 | 9 | 16 | 510 | 3.1 | 65 | 27 | 41 | 178 | 0.7 | 23 | 40 | 55 |
| **Jamaica** | 302 | 2.4 | 10 | 0 | 0 | 777 | 2.1 | 26 | 2 | 3 | 1294 | 3.2 | 44 | 12 | 16 | 389 | 1.1 | 13 | 21 | 26 |
| **St. Lucia** | 64 | 1.5 | 35 | 5 | 7 | 195 | 2.6 | 106 | 20 | 25 | 72 | 0.7 | 39 | 28 | 31 | 11 | 0.3 | 6 | 30 | 33 |
| **T&T<sup>D</sup>** | 125 | 1.8 | 9 | 0 | 0 | 961 | 3.1 | 68 | 7 | 13 | 2646 | 2.7 | 189 | 30 | 38 | 253 | 0.9 | 18 | 47 | 49 |
| **UK** | 54184 | 20.2 | 80 | 0 | 0 | 96692 | 2.3 | 144 | 7 | 28 | 34645 | 0.2 | 51 | 50 | 74 | 11965 | 0.3 | 18 | 86 | 92 |

<sup>A</sup> Crude mortality rate per 100,000 population
<sup>B</sup> Two or more doses of a 2-dose series or a single-dose COVID-19 vaccine; Average of vaccination coverage at start and end of waves calculated
<sup>C</sup> Any dose of a COVID-19 vaccine; Average of vaccination coverage with ≥1 dose of a COVID-19 vaccine at start and end of waves calculated
<sup>D</sup> T&T = Trinidad and Tobago
Sources: Our World in Data; UK Health Security Agency

Most Caribbean countries reviewed recorded their highest CFR and CMR during the third wave, with the highest CMR observed in T&T (189 per 100,000 population) and the highest CFR for Bahamas (4.1%). Exceptions were Barbados and St. Lucia where highest CMR occurred in their 2^nd^ wave (72 and 106 per 100,000 population respectively). In contrast, the UK experienced their highest CFR in the first wave (20.2%) with dramatic declines for subsequent waves (2.3% [2^nd^], 0.2% [3^rd^] and 0.3% [4^th^]). While the CFR for the UK was at least 10 times higher than the reviewed Caribbean countries in the first wave, UK’s CFR was significantly (at least 50%) lower than these Caribbean countries in the subsequent waves.

Vaccines only became available late in the first wave and just for Barbados and St Lucia. None of the Caribbean countries reviewed achieved the World Health Organization’s target of 70% vaccination by June 2022. At 12-Aug-2022, full vaccination coverage ranged from 25.5% (Jamaica) to 58.9% (Guyana); with less than half of the population fully vaccinated in three countries. The average full vaccination coverage during the fourth wave ranged from 21% in Jamaica to 54% in Barbados compared to 86% in the UK.

## Discussion

This analysis has shown that CFRs in the first wave of the COVID-19 pandemic was substantially lower in the Caribbean than in the UK, North America and other regions. Caribbean CFRs ranged from 1 to 3% in the first wave compared to 7% in Italy, 9-12 % in North America and 20% in South Africa.^20–22^ This was probably due to implementation of strong non-pharmaceutical measures including restricting travel into these countries early in the pandemic. Differences in age structure may account for CFR differences as the highest risk age group, persons 70 years and older, are more prevalent in HIC than LMIC.^23^ Population estimates indicate that persons 70 years and older account for 13% of the UK population but 7% in St. Lucia, Jamaica and T&T, and 5% in Guyana and The Bahamas. However, the SMR <1 between 29-Jun-2020 and 31-May-2021 in this analysis suggests that population age structure may not have been a major factor contributing to the lower first wave CFR observed in the Caribbean. In fact, Barbados, whose age structure is similar to the UK, with 12% of the population 70 years and older, was least affected by COVID-19 at this stage of the pandemic, even after adjustment for age (SMR 0.02), and fared better than the UK overall (SMR 0.62). Barbados implemented strict non-pharmaceutical measures and rolled out the COVID-19 vaccine early, targeting the elderly and most vulnerable first.

Case fatality rates and CMRs are influenced by COVID-19 case detection and reporting, testing coverage, vaccination coverage and access to treatment/appropriate management as well as inherent characteristics of the circulating VOC. While genotyping is limited in the Caribbean and country specific data on the dominant strains during each wave is not readily available, our analysis gives insight into the pattern of mortality for circulating VOC in the Caribbean. Among the Caribbean countries reviewed, Delta variant was first detected in Barbados (June 2021) then T&T, St. Lucia and Jamaica (August 2021); Bahamas (September 2021) and Guyana (December 2021). Prior to emergence of the Delta variant, Alpha and Gamma variants were the dominant circulating strains. Omicron dominated among all Caribbean Community (CARICOM) member states beyond January 2022.^24^ Therefore, generally the 1^st^ and 2^nd^ waves were associated with the Alpha variant while 3^rd^ and 4^th^ waves were associated with Delta and Omicron VOCs respectively.

The low SMRs observed in the Caribbean may partly be due to differences in case/death ascertainment across countries. Many Caribbean countries use the WHO definition for COVID-19 death (“death resulting from a clinically compatible illness, in a probable or confirmed COVID-19 case, unless there is a clear alternative cause of death that cannot be related to COVID disease, e.g. trauma).”^25^ The UKHSA however only records laboratory confirmed COVID-19 deaths. This difference in case definitions may inflate observed deaths and SMRs in the Caribbean. Underreporting of deaths (resulting in lower SMR) is also possible. Population density differences also were not correlated and ranged from a low 3.9 persons/square kilometre in Guyana to 666.1 in Barbados.^26^

The widespread implementation of non-pharmaceutical interventions (NPIs) for extended periods in the Caribbean is a more plausible explanation for the overall patterns of mortality during the first wave. Murphy et al ^27^ showed that NPIs in the Caribbean such as border closure, curfews, stay at home orders, and restrictions on gatherings occurred 4 to 30 days earlier in the Caribbean compared to international comparators, including the UK. A meta-analysis of NPIs during the pandemic showed that these interventions resulted in decreases in daily case growths (4.7% decline), daily death growth rates (4.8% decline), reproduction number (1.9% decline) and COVID-19 daily intensive care unit admissions (19.5% decline).^28^ Additionally, stay at home orders for at risk populations (e.g. persons with underlying illnesses and elders) continued up to the third wave (December 2021) in many Caribbean countries and this may have influenced the risk profile, and hence case fatality, of persons at highest risk of being affected by COVID-19.

The highest CMRs were observed in waves corresponding to detection of the Delta variant (third waves for Jamaica, Bahamas, Guyana, and T&T; second wave for Barbados, St. Lucia). Most countries experienced lower CMRs and CFRs in the fourth wave when the Omicron variant became the dominant strain after January 2022, with some protection provided by access to vaccines. This is consistent with global patterns of higher mortality risk for infection with Delta compared to other variants.^29^ In the USA the risk of COVID-19 death was 60% lower for infection with the Omicron variant compared to Delta variant. In-hospital mortality during the Delta wave was 15.1% compared to 4.9% during the first Omicron wave. ^30,31^ While some postulate that Omicron is a less virulent variant, higher levels of natural or acquired immunity (through vaccination) and changing demographics of the infected population (infection of younger persons during later waves) could account for lower mortality rates when Omicron emerged.

Although declines in CFR and CMRs were recorded in most countries after the third wave, the more dramatic decline in mortality rates for the UK occurred after the first wave. The transition to SMRs exceeding one for these Caribbean countries during subsequent waves emphasize global inequities in access to vaccination and novel therapeutics for COVID-19. Most Caribbean islands received their first set of COVID-19 vaccines between 15-Feb-2021 and 15-Mar-2021. In contrast, UK and United States of America (USA) began COVID-19 vaccination by Dec-2020.^32^ More than 50% of the doses of COVID-19 vaccination worldwide went to a handful of wealthy countries early in the pandemic. This inverse relationship between resources and COVID-19 mortality in later waves of the COVID-19 pandemic has been documented in other regions.^6^

Levels of vaccination against COVID-19 varied across the Caribbean during each wave. In most countries older persons were preferentially selected for vaccination early in the process, enhancing the role of vaccination in reducing mortality due to COVID-19. Barbados, a high-income country with the highest vaccination level of the countries in this analysis, experienced declines in CMR and CFR during second and third waves despite detection of the Delta variant during their second wave (Jun-2021). The increase in CMR during the fourth wave may reflect relaxation of non-pharmaceutical measures and waning immunity as Barbados was one of the first Caribbean islands to begin COVID-19 vaccination (Feb-2021). Jamaica, the third largest island in the Caribbean, had the second lowest vaccination coverage in the Caribbean during the third wave, with 12% of the Jamaican population fully vaccinated against COVID-19 compared to 28% in St. Lucia and 52% in Barbados. During the fourth wave, Jamaica had the lowest coverage.^33^

Despite low vaccination against COVID-19 in Jamaica, higher CMRs were observed in countries with better levels of vaccination during the third wave (Bahamas, T&T and Guyana). However, vaccination coverage in these Caribbean countries was well below the WHO target of 70% by mid-2022 and does not fully explain the differences within the Caribbean during waves. Differences between countries in vaccines type and effectiveness, duration of waves, compliance with NPI norms, risk profile of the affected population, access to therapeutics and resilience of health systems during surges may also contribute to these patterns in mortality rates. These factors need further exploration in order to understand the trends in mortality observed across the Caribbean. Regional sero-prevalence studies could also provide information on the role of natural immunity in later waves.

### Limitations

This analysis has several limitations. We estimated start and end dates of the waves. While inaccuracies with selection of dates are unlikely to affect the CFR, they have significant impact on the number of estimated deaths and hence the CMR. Many deaths associated with specific waves occur two to three weeks after a decline in cases and could contribute to underestimating the CMR for each wave. We verified start and end dates of waves by reviewing Ministry of Health, CARPHA reports/announcements and COVID-19 updates on local news media. As genetic sequencing is not widely available in the Caribbean, we applied regional data on dominant strains of COVID-19 to describe the association between VOC and mortality patterns in each country. It is possible that other VOCs accounted for high proportions of COVID-19 infections and deaths in specific countries that were not consistent with regional data.

Each country experienced successive waves at different time points and would be impacted by advancements in COVID-19 management at the time of the wave including availability of vaccination, access to new modalities of treatment and better understanding of disease management. While we accounted for vaccination coverage, other factors were not reliably documented.

Gaps in surveillance systems in the Caribbean also threaten the validity of this assessment. Additionally, rapid testing became more available after the third wave and could affect CFRs as persons with milder illness diagnosed by home testing were not captured by surveillance systems.

Up to the third wave, corticosteroids, which are inexpensive and widely available, emerged as an effective option for severe COVID-19 in clinical trials.^34,35^ However, access to other promising experimental treatments such as Remdesivir and monoclonal antibodies were not as widely available in the Caribbean compared to high income countries. These differences would also affect COVID-19 outcomes including mortality.

## Conclusion

The Caribbean experienced lower mortality due to COVID-19 during the initial stages of the pandemic (first wave) compared to the UK, probably due to implementation of various NPIs. However, limited access to COVID-19 vaccination and slow roll-out negatively impacted COVID-19 mortality during subsequent (second, third and fourth) waves. Generally, higher mortality was observed during circulation of the Delta variant, declining once the Omicron variant became dominant.

## Data Availability

All data produced are available online at Johns Hopkins Coronavirus Resource Center (https://coronavirus.jhu.edu/) and Our World in Data (https://ourworldindata.org/coronavirus).

## Acknowledgments

None

